# Primary human intestinal organoids recapitulate enteric infection of monkeypox virus and enable scalable drug discovery

**DOI:** 10.1101/2025.04.21.25325586

**Authors:** Pengfei Li, Jiangrong Zhou, Yining Wang, Xin Wang, Guige Xu, Rick Schraauwen, Ana Maria Gonçalves, Charlotte de Henau, Roberto Incitti, Dewy M. Offermans, Annemarie C. de Vries, Denis E. Kainov, Intikhab Alam, Karine Raymond, Amaro Nunes Duarte-Neto, Marcel J. C. Bijvelds, Qiuwei Pan

## Abstract

Monkeypox virus (MPXV) infection-associated intestinal manifestations including diarrhea and proctitis have been frequently reported during mpox outbreaks. The clade IIb MPXV strain has caused the 2022-2023 global outbreak, whereas the Ia and Ib strains are causing the concurrent outbreaks in Africa. Here, we found clinical evidence that MPXV can directly infect human intestine to induce lesions. Intriguingly, primary organoids cultured from human ileum and rectum support productive infections of MPXV clade IIb, Ia and Ib strains. Upon differentiation, we found that enterocytes and goblet cells but not enteroendocrine cells are capable of supporting viral replication. Given that primary intestinal organoids can be rapidly expanded in large scale, we were able to screen a broad-spectrum antiviral drug library. We identified 12 leading candidates of safe-in-human agents including clinically used drugs such as clofarabine. We extensively validated the anti-MPXV activity of clofarabine in human intestinal and skin organoids, and consistently demonstrated the potent antiviral activity against clade Ia, Ib and IIb strains. These findings are important for better understanding the clinical manifestations of mpox. Primary intestinal organoids-based infection models and the established antiviral drug discovery pipeline bear major implications in responding to the current mpox global health emergency, and sustaining epidemic poxvirus preparedness.

## Introduction

The monkeypox virus (MPXV) belongs to orthopoxvirus genus of the Poxviridae family. MPXV has a linear double-stranded DNA genome of about 200 kb in size, which encodes about 200 viral proteins (1). MPXV is comprised of two distinct clades: clade I with subclades Ia and Ib, and clade II with subclades IIa and IIb. The clade IIb strain has caused the 2022–2023 global outbreak of mpox (2), whereas the clade Ia and Ib strains are causing the concurrent outbreaks in Africa (3, 4). The World Health Organization (WHO) has two times declared a public health emergency of international concern (PHEIC) for the 2022–2023 global mpox outbreak and the concurrent outbreaks in Africa, respectively (5).

Although skin lesion is the most classical symptom, MPXV infection can cause a broad-spectrum systemic manifestations such as diarrhea, liver injury, myopericarditis, acute kidney injury and respiratory complications (6–8). Notably, proctitis—an inflammatory disorder of the rectum—has been diagnosed as a new clinical presentation during the 2022–2023 global mpox outbreak, with an incidence rate of about 20% (9, 10). In mpox patients with gastrointestinal manifestations, proctitis and symptoms of rectal pain, diarrhea, and vomiting are prevalent (11). Systemic manifestations are often associated with worse clinical outcomes (8, 9), but such atypical presentations of mpox remain poorly studied.

Patients with severe mpox require hospitalization, supportive care and antiviral treatment, but no approved medication is available for specifically treating MPXV infection. The antiviral drug tecovirimat was approved for treating smallpox under the US FDA’s Animal Rule, which is based on its efficacy in relevant animal models using related orthopoxviruses including MPXV (12). It blocks the final steps in virus maturation and release from the infected cell by disrupting the major envelope wrapping protein VP37 (13). Tecovirimat has been widely prescribed as compassionate use for treating clade IIb MPXV infection during the global outbreak (14). However, results from two recently conducted randomized, placebo-controlled trials showed no clinical benefits of tecovirimat treatment for children and adults infected with clade I MPXV (15), or adults infected with clade II MPXV (16). Cidofovir and its prodrug brincidofovir, functioning as viral DNA polymerase inhibitors (13), have also been occasionally prescribed for treating mpox, but their clinical efficacy remains undefined (17).

Giving the common prevalence of mpox-associated intestinal manifestations, we first probed the clinical evidence and found that MPXV can directly infect the human intestine and cause numerous lesions. Next, we demonstrated that primary organoids derived from human small intestine and rectal tissues support productive infection of both clade I and II MPXV isolates. The infection triggered robust virus-host interactions. Since primary intestinal organoids can be expanded in large scale, we conducted antiviral drug screening and identified potent inhibitors against MPXV infection.

## Results

### Clade IIb MPXV infection in human intestine and cultured primary intestinal organoids

Intestinal manifestations in particular proctitis have been frequently reported during the 2022-2023 global outbreak, which was caused by the clade IIb MPXV strain (9, 10). To investigate whether MPXV can directly infect the intestinal tract, we examined colon tissue from our previously reported fatal mpox case (Figure 1A) (18). In this autopsied mpox patient, we observed numerous lesions in the colon (Figure 1B). H&E staining of the descending colon showed intestinal gland with degenerated goblet cells characterized by shrunken, eosinophilic cytoplasms and condensed nuclear chromatin, alongside exocytosis of inflammatory cells (Figure 1C). The lamina propria exhibited inflammatory cells with eosinophilic cytoplasms, corresponding to a Guarnieri-like inclusion, and nuclear alterations. A small venule in the mucosa shows luminal inflammatory cells and an endothelial cell containing a Guarnieri-like inclusion in the cytoplasm (Figure 1C). Immunohistochemistry staining of the viral antigens and counterstained with Alcian blue for mucus in goblet cells revealed numerous infected cells in the lamina propria as well as in the goblet cells, with intestinal glands partially or entirely positive for the viral antigens (Figure 1D). Furthermore, colonic epithelium (CDX2 positive) was infected by MPXV, with cytopathogenic effects including evidence of apoptosis and necrosis of epithelial cells (Figure 1E-G; Supplementary Figure S1). IHC staining to the intestinal wall near peritoneum showed massive MPXV-positive cells, and interestingly, MPXV infection was also observed in endothelial cells and interstitial intestinal cells (Figure 1H). These results prompted us to investigate whether human intestinal organoids are permissive to clade IIb MPXV infection, by using a patient-derived isolate from the 2022 outbreak in the Netherlands.

**Figure 1.**
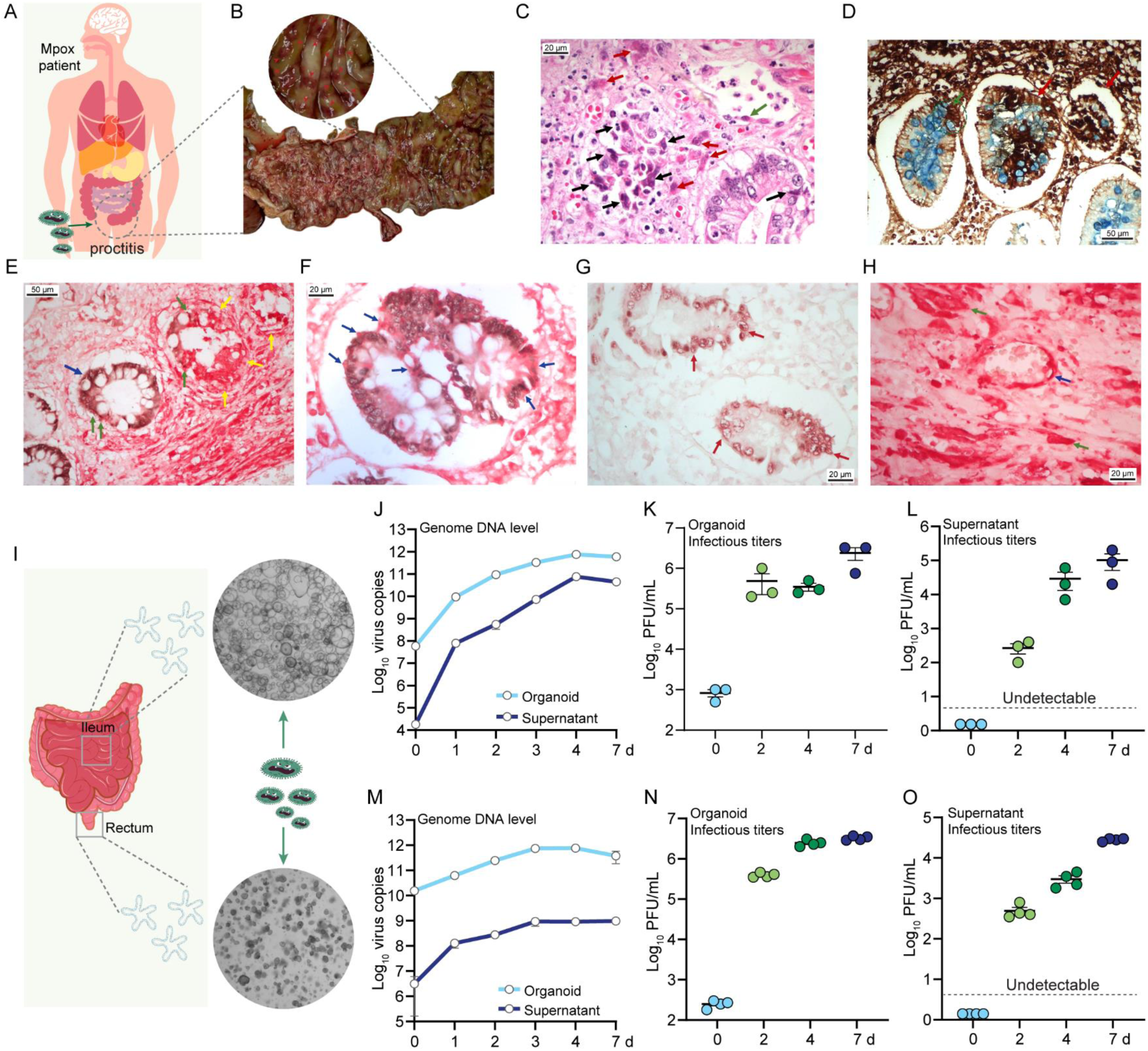
Clade IIb MPXV infection in human intestine and cultured primary intestinal organoids. (A) Schematic diagram of MPXV infection in human intestine (colon). (B) MPXV induced severe lesions in descending colon tissue of the infected patient (Red arrows indicate the lesions). (C) H&E staining of MPXV infected descending colon. Black arrows indicate shrunken and eosinophilic cytoplasms, and condensed nuclear chromatin in cells; red arrows indicate guarnieri- like inclusion and nuclear alterations; green arrow indicates the guarnieri-like inclusion in the cytoplasm of an endothelial cell. (D) Immunohistochemistry staining with anti-vaccinia antibody (for MPXV) and Alcian blue staining (for mucus in goblet cells) in descending colon tissue. Green arrow indicates an intestinal gland with partially staining of viral antigen, and red arrows indicate almost the entire gland of positive staining. (E) IHC staining with anti-vaccinia antibody (red; detecting MPXV virions) and anti-CDX2 (brown) in intestinal mucosa with ulcerative lesions. Blue arrow indicates increased nuclei and vacuolated cytoplasm in the epithelium. Green arrows indicate apoptotic epithelial cells with pycnotic nuclei induced by MPXV infection. Yellow arrows indicate MPXV-positive and necrotic epithelial cells. (F) Magnified image of IHC staining (anti-vaccinia, red; anti-CDX2, brown) in intestinal mucosa with ulcerative lesions. Blue arrows indicate that the epithelium is reactive, with increased nuclei and vacuolated cytoplasm by MPXV infection. (G) IHC staining of anti-vaccinia (red) and anti-CDX2 (brown) in intestinal mucosa distant from ulcerative lesions. Red arrows indicate the epithelium with increased nuclei and lamina propria with edema. Anti-CDX2 is positive and anti-vaccinia signal is negative. (H) IHC staining with anti-vaccinia antibody (red) to the intestinal wall near peritoneum. Blue arrow indicates endothelial cells in an intestinal small vessel. Green arrows indicate MPXV infection in interstitial intestinal cells. (I) Schematic representation of primary intestinal organoids isolation, and the bright field image of organoids-derived from ileum and rectum tissues. (J) Quantification of viral DNA levels from ileum organoids or culture medium (0 in x axis represent 1 hour post-infection; n= 4). (K) Quantification of infectious viral titers of intracellular virus in ileum organoids (n= 3). (L) Quantification of infectious viral titers of ileum organoid secreted viruses in medium (n= 3). (M) Quantification of viral DNA levels from rectum organoids or culture medium (n= 3). (N) Quantification of infectious viral titers of intracellular virus in rectum organoids (n= 4). (O) Quantification of infectious viral titers of rectum organoid secreted viruses in medium (n= 4).

Firstly, we inoculated the primary organoids isolated from human small intestine (ileum) (Figure 1I). By qRT-PCR quantification the viral DNA level, we observed a continuous increase of intracellular viral genome copies from 1 hour, 1 day and up to 7 days post-inoculation, and the kinetics of extracellular virus secreted into culture medium showed a similar increasing trend (Figure 1J). In organoids, plaque assay revealed that the infectious titers increased from approximately 3 log10 plaque-forming units (PFU)/ml at 1 hour to nearly 6.5 log10 PFU/ml at 7 days post-inoculation (Figure 1K). In culture medium, the infectious titer was undetectable at 1 hour and peaked at nearly 5 log10 PFU/mL at 7 days post-inoculation (Figure 1L). Next, we tested human rectum tissue-isolated organoids (Figure 1I), and consistently, continuous viral replication in organoids and production into medium was observed for both viral DNA level and infectious titers (Figure 1M-O).

Next, the robust infection was further visualized by immunostaining MPXV virions in both types of organoids (Figure 2A and 2B). Primary intestinal organoids typically exhibit features of both epithelial cells (EpCAM+) and stem cells (SOX9+), along with robust proliferative capacity (Ki67+; Figure 2C). We thus co-stained MPXV with these markers. We observed that MPXV can infect both proliferating cells and stem cells (Figure 2D and 2E). We specifically included intestinal epithelium marker CDX2, and in line with our clinical findings (Figure 1E and 1F), we found MPXV infection in CDX2-positive organoid cells (Figure 2F). In addition, infected organoids exhibited marked morphological shrinkage after 7 days of infection, and immunostaining revealed disruption of tight junctions (ZO-1+) and widespread apoptotic cell death (cleaved caspase-3+) in these organoids (Figure 1G; Supplementary Figure S2). Transmission electron microscopy (TEM) visualized the intracellular MPXV particles, with the majority captured at the immature and mature virion stages in intestinal organoids (Figure 2H and 2I).

**Figure 2.**
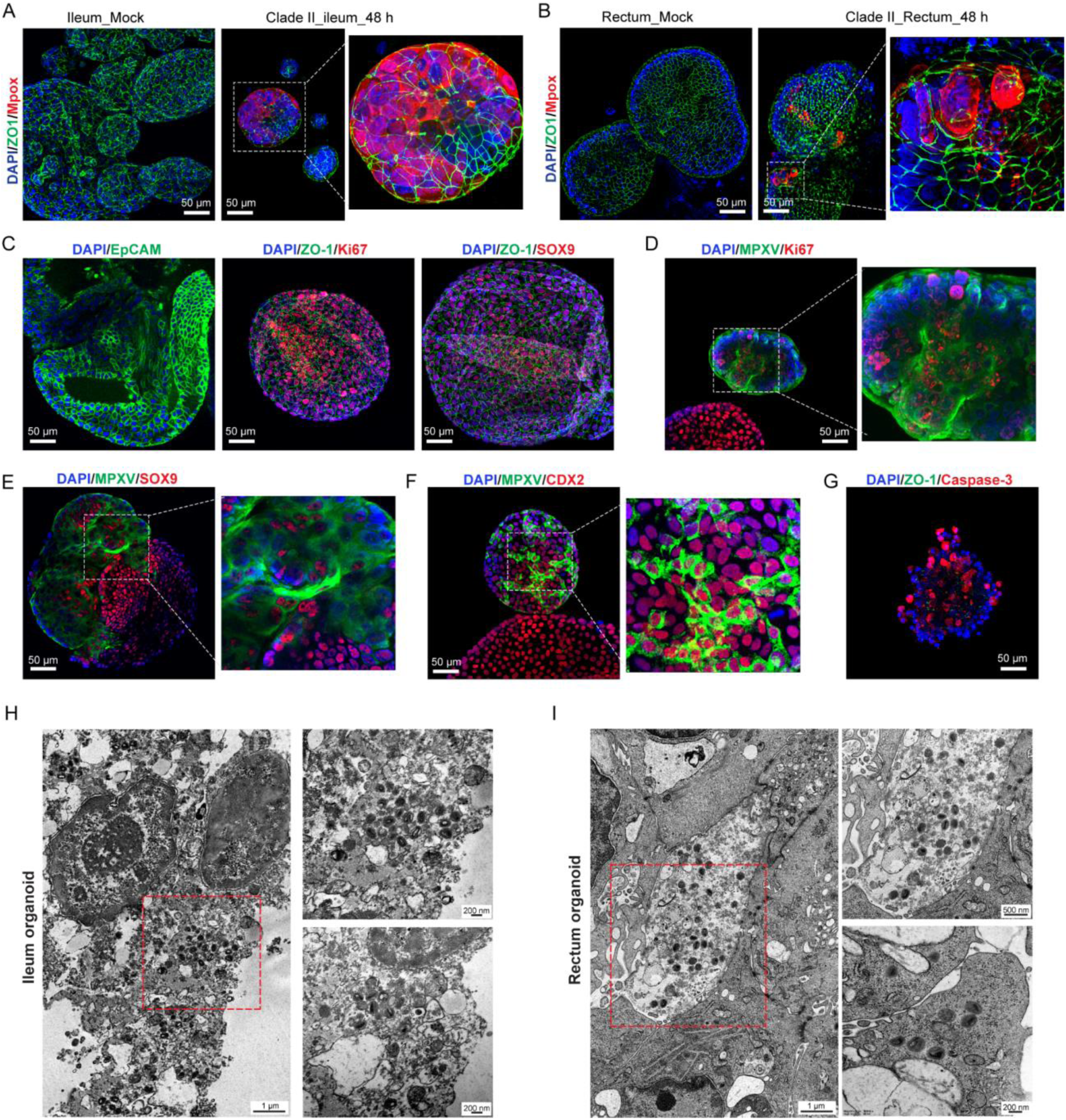
Visualization of MPXV infection in intestinal organoids. Visualization of MPXV-infected ileum organoids (A) and rectum organoids (B) by immunostaining with antibodies against ZO-1 (green), MPXV virions (red), and DAPI nuclei staining (blue). (C) Characterization of intestinal organoids by staining epithelial membrane marker EpCAM, tight-junction marker ZO-1, proliferative marker Ki67 and stem cell marker SOX9. Co-staining MPXV with Ki67 (D), SOX9 (E), and intestinal epithelium marker CDX2 (F) at 48 hours post-infection. (G) Immunostaining cell death marker cleaved caspase-3 and tight-junction marker ZO-1 at 7 days post-infection. Representative TEM visualized MPXV virions in ileum (H) and rectum (I) organoids.

### The differential susceptibility of different types of enteric epithelial cells to MPXV infection

To further investigate the infectivity of MPXV to different types of enteric epithelial cells, intestinal organoids were further differentiated towards enterocytes, goblet cells and enteroendocrine cells (Figure 3A and 3B; Supplementary Figure S3A). These differentiated organoids were subsequently inoculated with clade IIb MPXV particles, and qRT-PCR quantification of viral DNA indicated that goblet cells- and enterocytes-differentiated organoids were readily supportive to MPXV replication and production (Figure 3C and 3D). Immunostaining of MPXV virions further confirmed MPXV infections in goblet cells- and enterocytes-differentiated organoids (Figure 3E and 3F). However, enteroendocrine-differentiated organoids appeared to not support MPXV replication, showing decreased viral DNA levels at 48 hours and undetectable MPXV fluorescence signal in CHGA-positive cells (Figure 3G; Supplementary Figure S3B). In addition, we used ileal organoids-derived cells to grow epithelial monolayers on a permeable (trans-well) support. Upon viral inoculation from the apical (luminal) compartment (Figure 3H), we observed robust viral infections by immunostaining at 48 hours post-inoculation (Figure 3I). QRT-PCR quantification showed viral secretion into both the apical (up to 11 log_10_ copies) and the basolateral (up to 7 log_10_ copies) compartments, although over 99% of the total production was in the apical compartment (Figure 3J).

**Figure 3.**
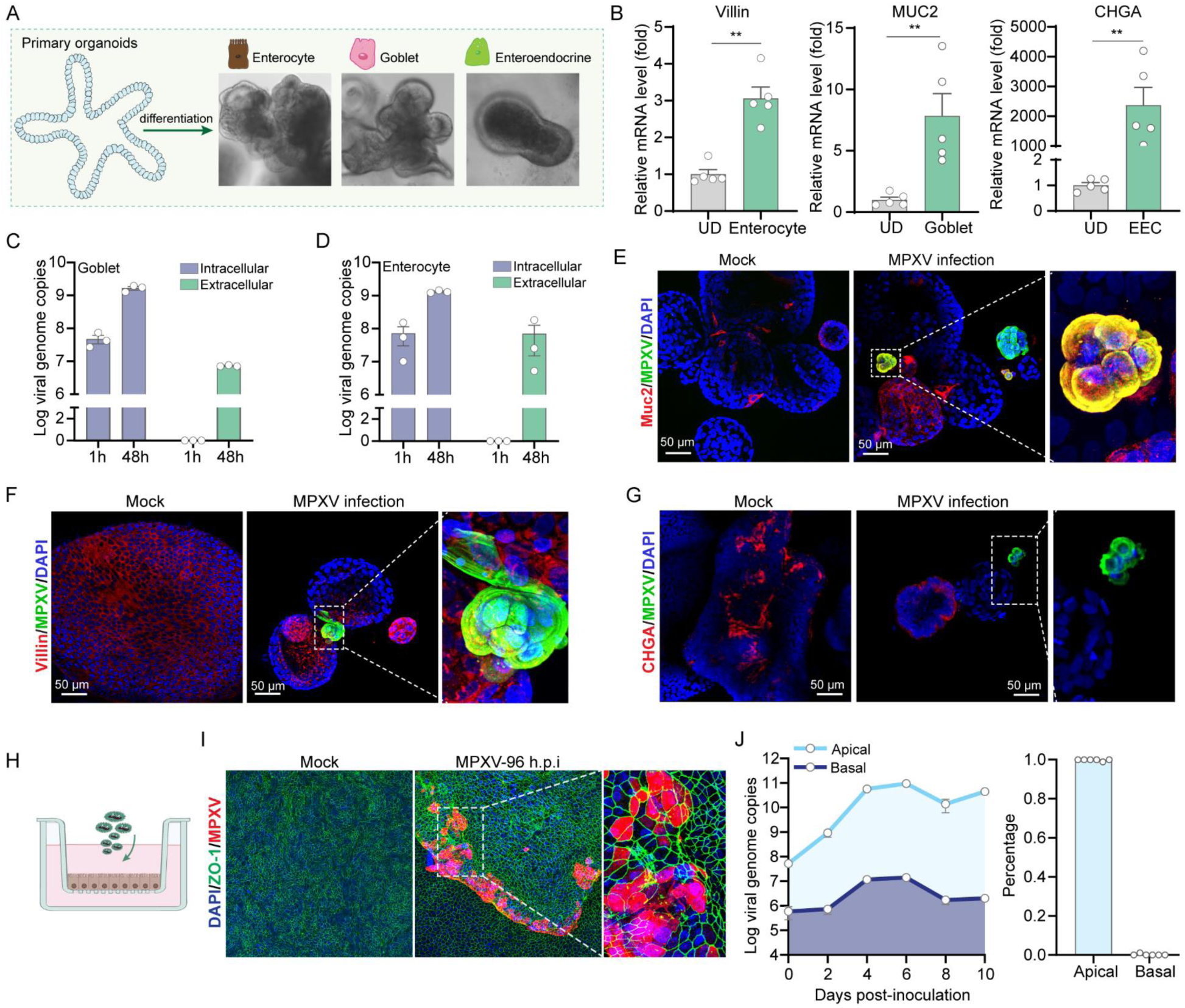
MPXV infection in differentiated intestinal organoids. (A) Morphology of intestinal organoids upon differentiation culture towards enterocyte-, goblet-, and enteroendocrine-phenotype. (B) Gene expression level of Villin (enterocytes marker), Muc2 (goblet cell marker) and CHGA (enteroendocrine cell marker) upon different protocols of differentiation culture. UD: undifferentiated organoids; EEC: enteroendocrine-differentiated organoids. Data are presented as mean ± SEM. Statistical analysis was performed using the two-tailed Mann–Whitney test. \*\**P < 0.01*. Quantification of viral DNA levels from goblet-differentiated organoids and its culture medium (C), as well as from enterocyte-differentiated organoids and culture medium (D) at 48 hours post-infection (n = 3). Immunofluorescence staining of goblet-differentiated (E), enterocyte-differentiated (F), and enteroendocrine-differentiated (G) organoids with MPXV infections for 48 hours. Muc2 (goblet cells, red), Villin (enterocytes, red), CHGA (enteroendocrine cells, red). In enteroendocrine-differentiated organoids, the rare MPXV signals were only found in CHGA-negative cells. (H) Schematic diagram of MPXV infection in trans-well cultured organoid cell monolayers. (I) Immunostaining of ZO-1 (green), MPXV (red) and nuclei (DAPI, blue) in organoid cell monolayers at 96 hours post-infection. (J) Quantification of released viruses from apical or basolateral compartment of trans-well system.

### Transcriptomic analyses reveal active MPXV– host interactions

As a large DNA virus, MPXV is supposed to transcribe hundreds of viral genes during infection (19). We performed transcriptomic analysis on intestinal organoids infected with clade IIb MPXV (Figure 4A), and notably, MPXV transcript levels were identified to increase over the 96 hours post-inoculation (h.p.i.) gradually (Figure 4B). In-depth analysis revealed a number of abundantly expressed transcripts mapped to the different locations of the MPXV reference genome in a temporal expression patterns (Figure 4C).

**Figure 4.**
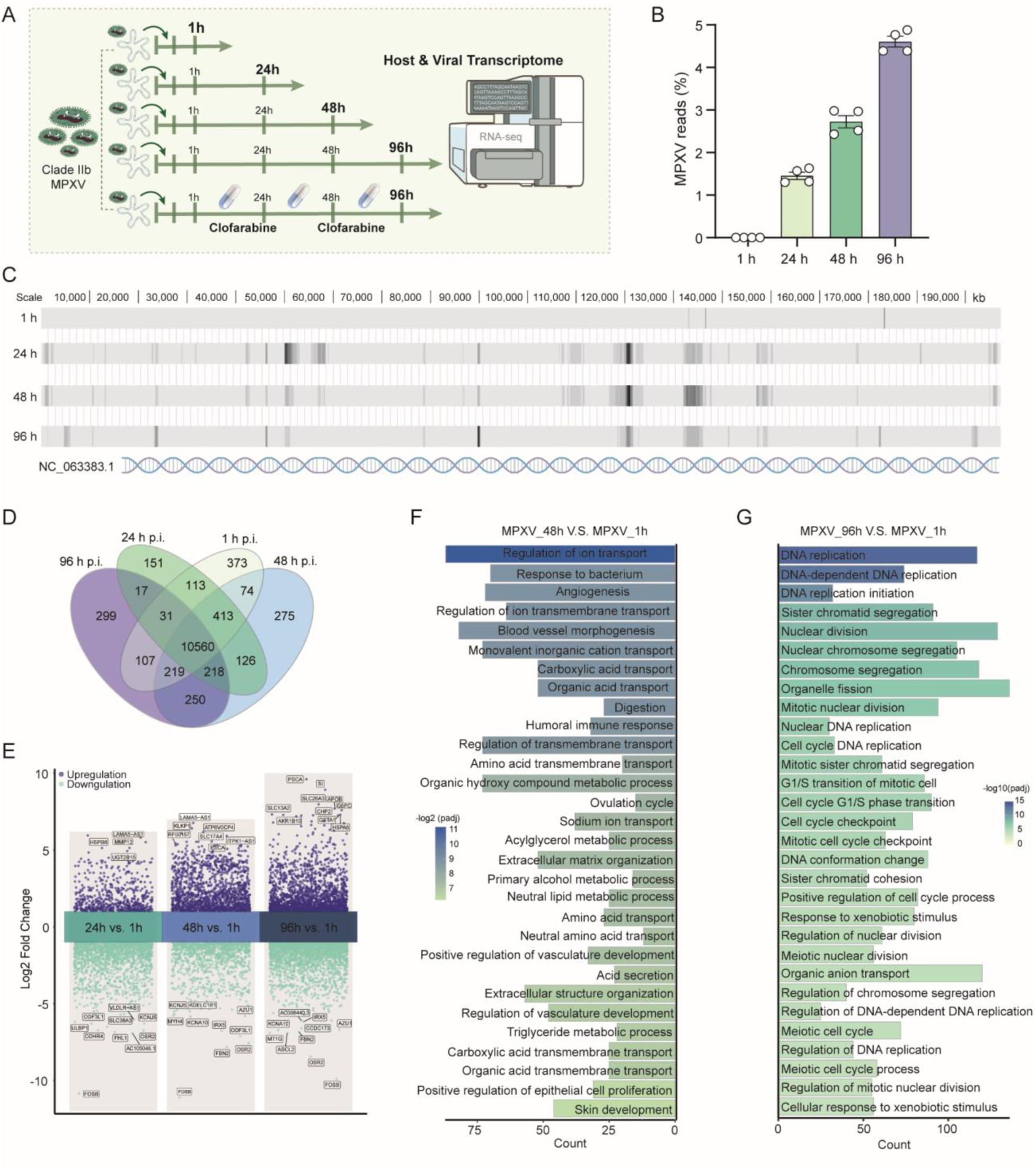
Mapping host and virus transcriptomes. (A) Schematic representation of bulk-RNA sequencing groups. (B) The percentages of mapped MPXV transcripts in different groups of organoids. (C) MPXV transcripts mapped to the locations in viral genome. (D) Venn diagram of overlapped differentially expressed genes in MPXV infected organoids at different time points. (E) Volcano plot analysis of differentially expressed genes at 24, 48 and 96 hours compared to 1 hour post MPXV infection. (F, G) Top 30 significantly enriched pathways by gene ontology (GO) analysis of MPXV infected organoids at 48 hours and 96 hours, compared to 1 hour post-infection.

Comparison of the differentially expressed genes between different time points post-inoculation showed that hundreds of genes were uniquely expressed at each time point, while over 10,000 genes were consistently expressed across all time points (Figure 4D). Volcano plot analysis revealed robust host responses to MPXV infection at 24, 48, and 96 hours compared to 1 hour, with thousands of genes significantly up- or downregulated (Figure 4E). Intriguingly, several prominently regulated genes such as OSR2, FOSB, and PSCA, were consistently dysregulated by MPXV from 24 to 96 hours. Gene Ontology analysis revealed that at 48 h.p.i., pathways related to ion transport and immune response were significantly enriched (Figure 4F), whereas at 96 h.p.i., many significantly regulated pathways were associated with DNA replication compared to 1 h.p.i. (Figure 4G). In addition, transcriptional signatures related to unfolded protein responses and heat stress responses were enriched in infected organoids at 96 h.p.i. compared to un-infected controls (Supplementary Figure 4A-C). Overall, these findings demonstrate that MPXV, with its large DNA genome encoding numerous viral proteins, elicits specific and dynamic host responses in intestinal organoids throughout the course of infection.

### Drug screening in intestinal organoids identified leading candidates against clade IIb MPXV

Currently, effective antiviral treatments against MPXV infection are urgently needed (20). A unique advantage of primary intestinal organoids is their capability for rapid and large-scale expansion (21). This enabled us to screen a library of 240 known safe-in-human broad-spectrum antiviral (BSA) agents in clade IIb MPXV infected intestinal organoids (Figure 5A). In this study, we employed a low concentration (1 μM) to minimize nonspecific effects and cidofovir was included as a positive control. After treatment of 48 hours, 12 compounds were identified with over 80% inhibition on intracellular MPXV genome DNA (Figure 5B), which are equivalent or more potent than cidofovir. These 12 inhibitors were further categorized into four groups based on their original applications, including heart failure treatment group, leukemia treatment group, protein synthesis blocker group, and nucleoside analogue group (Figure 5C). Importantly, qRT-PCR quantification demonstrated that these 12 compounds can potently and dose-dependently inhibit the intracellular MPXV genomic DNA levels (Supplementary Figure 5A-F). Immunostaining of MPXV virions further demonstrated the potent inhibitory effect of these agents (Figure 5D). In addition, the anti-MPXV activity of these compounds was validated in human rectal organoids (Supplementary Figure 5G).

**Figure 5.**
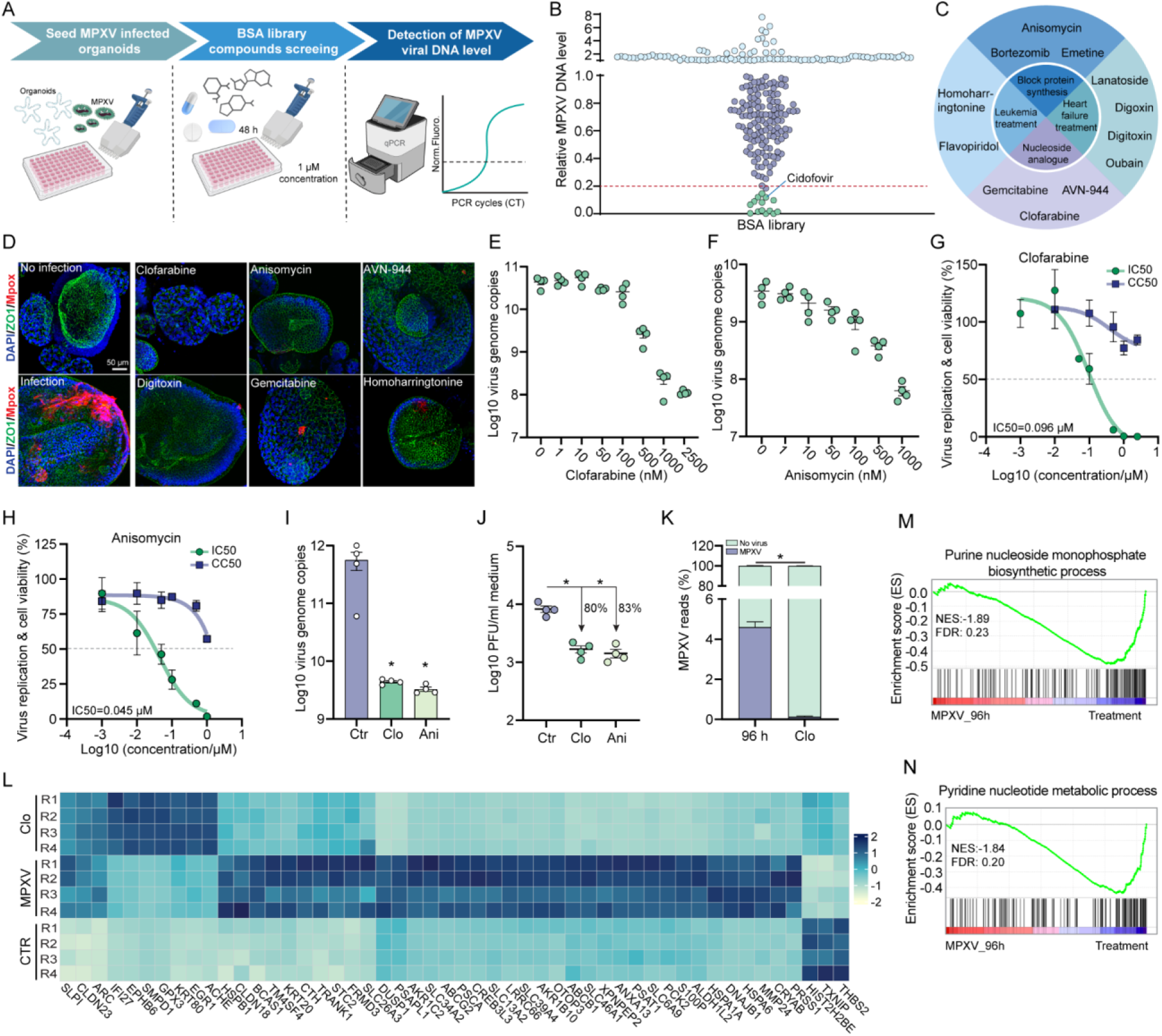
Antiviral drug discovery and validation in MPXV infected intestinal organoids. (A) Schematic diagram of broad-spectrum antiviral (BSA) library screening in MPXV infected intestinal organoids. (B) Quantification of MPXV DNA levels after treating with the 240 compounds in BSA library for 48 hours. (C) General information of the 12 identified potent MPXV inhibitors. (D) The inhibitory effect of 6 prioritized compounds by immunostaining MPXV virions in intestinal organoids at 48 h.p.i. The inhibitory activity of clofarabine (E) and anisomycin (F) against MPXV viral DNA in intestinal organoids treated for 48 hours. Half maximum inhibitory concentration (IC50) and half maximum cytotoxic concentration (CC50) of clofarabine (G) and anisomycin (H) in intestinal organoids infected with MPXV for 48 hours. (I) The inhibitory efficacy of clofarabine (Clo) and anisomycin (Ani) to intracellular MPXV DNA levels in intestinal organoids at the scenario of delayed treatment (the treatment initiated at 3 d.p.i). (J) Quantification of MPXV infectious titers in culture medium by 48 hours treatment in the scenario of delayed treatment (n= 4). (K) The percentages of mapped MPXV transcripts in organoids with 1 μM of clofarabine treatment or non-treatment after 96 hours infeciton (n = 4, *P < 0.05* by χ^2^ test). (L) Top 50 significantly regulated genes upon MPXV infection for 96 hours. (M-N) Gene set enrichment analysis (GSEA) including purine nucleoside monophosphate biosynthetic process (M) and pyridine nucleotide metabolic process (N) in intestinal organoids infected with MPXV alone compared to MPXV infection along with clofarabine treatment.

Notably, clofarabine and anisomycin, represented the most potent inhibitors of MPXV infection, reducing approximately 2.5 log_10_ and 1.8 log_10_ of MPXV DNA copies, respectively (Figure 5E and 5F). The estimated half maximal inhibitory inhibition (IC50) concentration of clofarabine and anisomycin was 0.096 and 0.045 μM respectively, with no major cytotoxicity at tested concentrations of both compounds (Figure 5G and 5H; Supplementary Figure 5H and 5I). In the clinic, there can often be a delay in receiving antiviral treatment for mpox patients (22). We then tested a scenario of delayed antiviral treatment. We initiated a 48 hours-treatment with 1 μM of clofarabine or anisomycin at 3 days post-infection. Quantification of intracellular MPXV DNA showed nearly 2 log10 reduction in viral DNA copies (Figure 5I). Plaque assay demonstrated significant inhibition of infectious virus production in culture medium (Figure 5J). We then modeled a relatively long-term treatment, with immediate treatment after viral inoculation and extended the treatment duration for 7 days. We observed significant inhibition of both intracellular viral replication and extracellular virus production (supplementary Figure 5J and 5K).

### Validation of clofarabine as a potent inhibitor against clade IIb MPXV in intestinal and skin organoids

Next, we prioritized clofarabine for further validation considering its favorable pharmacological profiles, and the fact that it is already FDA-approved for treating acute lymphoblastic leukaemia (23). By genome-wide transcriptomic analysis, we observed a dramatic reduction of MPXV transcripts in clofarabine-treated intestinal organoids compared with the untreated organoids at 96 hours (4.61% versus 0.14%; 97% inhibition) (Figure 5K). Differential gene expression analysis showed that the impact of rewiring host transcriptome by MPXV infection for 96 hours was largely prevented by clofarabine treatment (Figure 5L). Interestingly, gene set enrichment analysis revealed that transcriptional signatures associated with nucleoside biosynthetic processes were largely prevented by clofarabine treatment (Figure 5M and 5N; Supplementary Figure 4D-G).

Recently, we have developed MPXV infection models using human induced pluripotent stem cell-derived skin organoids (24). Here, we further validated the antiviral activity of clofarabine in air-liquid interface (ALI)-cultured skin organoids (Figure 6A). We first assessed the antiviral effect of immediate treatments over 7 days after viral inoculation. This resulted in a significant inhibition of virus production in culture medium at day 7 (Figure 6B and 6C). Correspondingly, infectious titers of produced virus were potently inhibited, with 93% reduction shown by plaque assay (Figure 6D). Consistently, intracellular virus replication was potently inhibited as demonstrated by quantifying viral DNA in organoids after 7 days of treatment (Figure 6E). Next, we performed a delayed treatment in cystic skin organoids, in which the treatment with clofarabine (1 μM) was initiated 4 days post-infection (Figure 6F). Quantification of MPXV DNA levels in culture medium showed a persistent inhibition of virus production from day 6 to 12 post-infection (Figure 6G).

**Figure 6.**
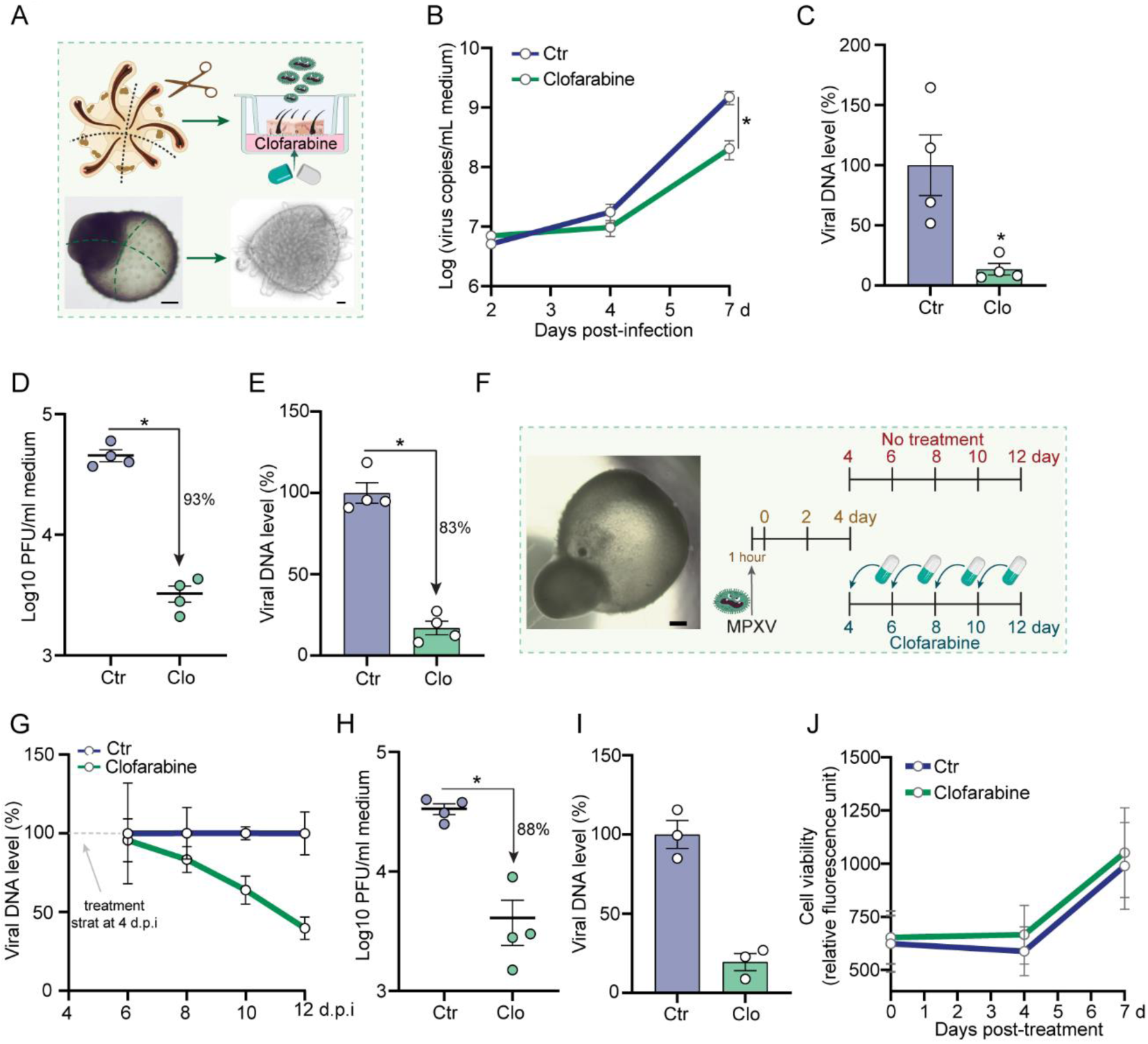
Validation of the anti-MPXV activity of clofarabine in human skin organoids. (A) Schematic illustration of ALI-cultured skin organoids. (B) Quantification of MPXV DNA level in culture medium of ALI-cultured skin organoids at 2, 4, and 7 days post-infection (n = 4). (C) Relative viral genome DNA level at 7 days post-infection (n = 4). (D) Quantification of MPXV infectious titers in culture medium at 7 days post-infection (n = 4). (E) Quantification of MPXV DNA level in skin organoids at 7 days post-infection (n = 4). (F) Schematic representation of delayed clofarabine treatment in skin organoids. (G) Quantification of MPXV DNA level in culture medium at 6 days, 8 days, 10 days, 12 days post-inoculation. Clofarabine (1 μM) was administered at 4 days post-inoculation (n = 4). (H) Quantification of infectious titers in the culture medium of skin organoids at 12 days post-inoculation (n = 4). (I) Quantification of MPXV DNA level in skin organoids at 12 days post-inoculation (n = 3). h.p.i: hour post-infection; d.p.i: day post-infection. (J) Cell viability of skin organoids with or without clofarabine treatment (1 μM) (n = 5).

On day 12, the treatment resulted in a 88% reduction of infectious virus titers (Figure 6H). Likewise, intracellular virus replication was potently inhibited as demonstrated by quantifying viral DNA in skin organoids on day 12 post-infection (Figure 6I). Importantly, cell viability assay showed that 1 μM of clofarabine treatment had no clear cytotoxicity to skin organoids (Figure 6J).

### Clofarabine inhibits clade Ia and Ib MPXV infections in intestinal organoids

Currently, mpox outbreaks caused by the clade Ia and Ib strains are devastating in Central and East Africa (3). We thus aimed to ascertain whether clofarabine can also suppress the replication of these strains. To this end, we inoculated intestinal organoids with patient derived clade Ia and Ib MPXV isolates (Figure 7A). We first tested the clade Ia isolate and observed a continuous increase of both intracellular and extracellular viral genome copies from 1, 24, and up to 96 h.p.i. (Figure 7B). Immunofluorescence staining further confirmed robust MPXV infections at 48 h.p.i. (Figure 7C). Similarly, intestinal organoids effectively support the replication and production of clade Ib MPXV, as demonstrated by qPCR quantification of the viral genome DNA and immunostaining of MPXV virions (Figure 7D and 7E).

**Figure 7.**
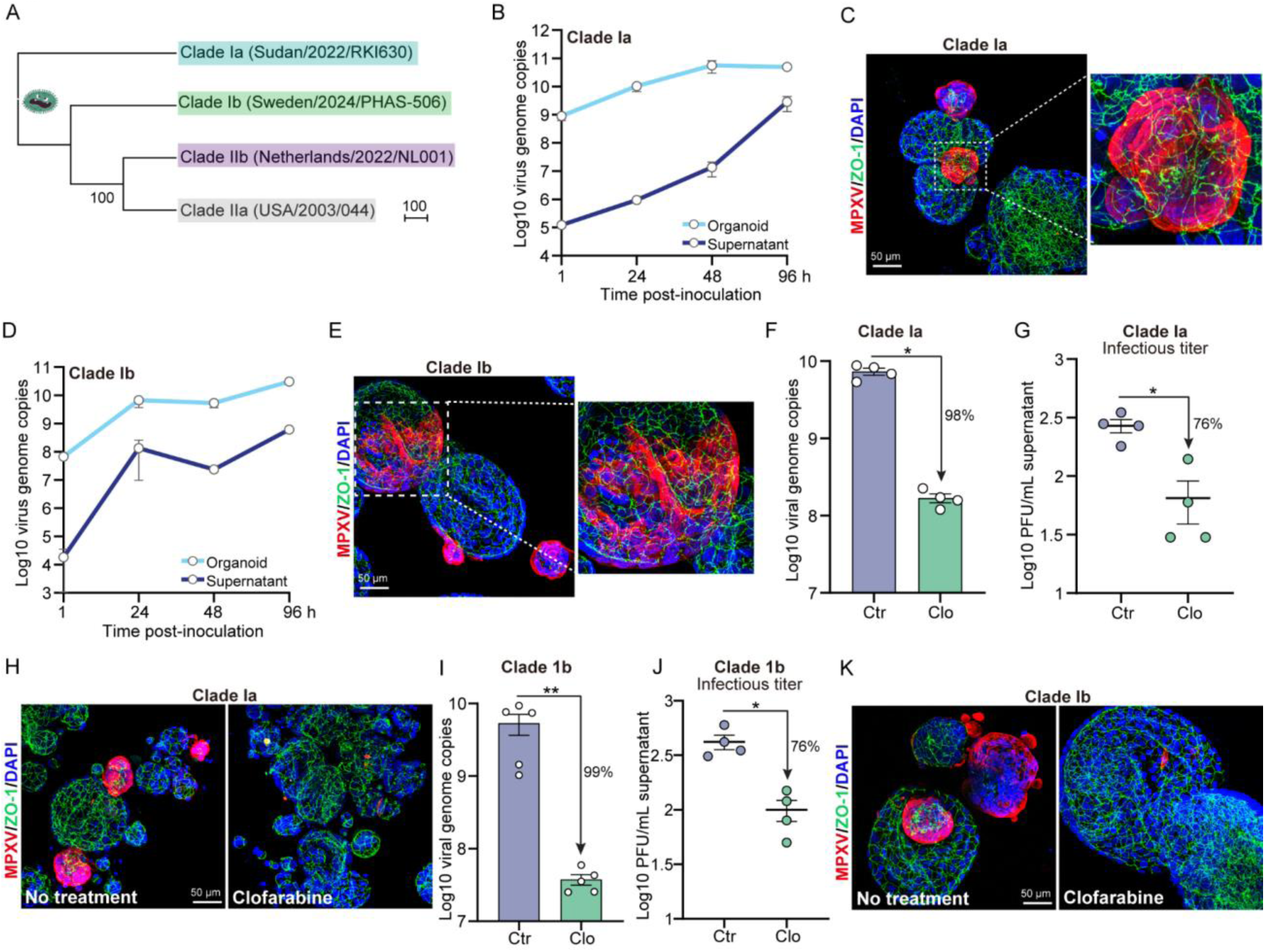
Recapitulating clade Ia and Ib MPXV infections and validating the antiviral effects of clofarabine in intestinal organoids. (A) Phylogenetic tree of clade I and II MPXV strains. (B) Quantification of viral DNA levels in intestinal organoids and culture medium at 1, 24, 48 and 96 hours post-infection with clade Ia MPXV strain (n = 4). (C) Representative immunostaining with antibodies against MPXV virions (red), ZO-1 (red) and DAPI nuclei staining (blue) in clade Ia MPXV infected intestinal organoids. (D) Quantification of viral DNA levels in intestinal organoids and culture medium at 1, 24, 48 and 96 hours post-infection with clade Ib MPXV strain (n = 4-5). (E) Representative immunostaining with antibodies against MPXV virions (red), ZO-1 (red) and DAPI nuclei staining (blue) in clade Ib MPXV infected intestinal organoids. (F) Quantification of the intracellular DNA level in intestinal organoids infected with clade Ia MPXV at 48 h.p.i (n = 4). (G) Quantification of the infectious virus titers in culture medium of clade Ia infected intestinal organoids at 48 h.p.i (n = 4). (H) Representative immunostaining with antibodies against MPXV virions (red), ZO-1 (red) and DAPI nuclei staining (blue) in clofarabine treated or non-treated intestinal organoids at 48 hours post infection of clade Ia strain. (I) Quantification of the viral DNA level in intestinal organoids infected with clade Ib MPXV at 48 h.p.i (n= 5). (J) Quantification of the infectious virus titers in culture medium of clade Ib infected intestinal organoids at 48 h.p.i (n = 4). (K) Immunostaining with antibodies against MPXV virions (red), ZO-1 (red) and DAPI nuclei staining (blue) in clofarabine treated or non-treated intestinal organoids at 48 hours post infection of clade Ib strain. h.p.i: hour post-infection. Data are presented as mean ± SEM. Statistical analysis was performed using the two-tailed Mann–Whitney test. \**P < 0.05*, \*\**P < 0.01*.

Lastly, we treated clade Ia or Ib infected intestinal organoids with 1 μM clofarabine. For the clade Ia MPXV strain, treatment of clofarabine for 48 hours inhibited 98% of intracellular viral DNA replication, and reduced infectious virus production in the culture medium by 76% (Figure 7F and 7G). Immunostaining further confirmed this potent inhibitory effect to clade Ia MPXV (Figure 7H). Consistently, clofarabine was capable of inhibiting clade Ib MPXV intracellular replication and extracellular production, as demonstrated by quantification of viral genome DNA, infectious virus titers and immunostaining the virions (Figure 7I-K). Collectively, clofarabine is effective against clade Ia and Ib MPXV.

## Discussion

In general, the common symptoms of mpox such as skin rash or mucosal lesions typically improve within a few weeks. However, some patients can experience disseminated diseases resulting in severe complications and even death. Histopathological evaluation of autopsy and biopsy tissues from fatal mpox patients who displayed systemic manifestations, showed that MPXV antigen and viral DNA were present in a wide range of tissues, including skin, ocular, oropharyngeal, and mucosal digestive tract tissues (25). In our autopsied mpox patient (18), we observed robust MPXV infection and the massive presence of lesions in the colon.

In line with these clinical observations, we demonstrated that primary organoids cultured from human small intestine and rectum are highly susceptible to MPXV infection. Furthermore, our model recapitulates MPXV-caused cell death, in line with the massive tissue damage observed in the intestine of our mpox patient. These organoids derived from tissue stem cells and cultured in 3D structure are capable of recapitulating the architecture, composition, diversity, organization, and functionality of cell types of the original intestinal tissue (26). We further differentiated intestinal organoids into three types of enteric cells. Interestingly, enterocytes and goblet cells but not enteroendocrine cells support MPXV replication. A recent study using human induced pluripotent stem cell-derived colon organoids failed to model robust infection of MPXV (27). Although the underlying reason remains unknown, we question whether their model lacks the cell types that are permissive to MPXV replication. When our organoids were grown into epithelial monolayers on a trans-well system in 2D, MPXV was predominantly secreted into the apical compartment, although basolateral secretion also occurred. This may reflect gastrointestinal shedding of MPXV, since the virus has been detected in stool samples from a proportion of mpox patients (28).

A unique advantage of primary organoids is capable of rapid expansion, and therefore enable small- to medium-scale drug screening (29). In this study, we focused on broad-spectrum antivirals (BSAs) that can inhibit the infection of multiple viruses from the same or different viral families by acting on conserved viral components or host cellular pathways shared by multiple viruses (30). This broad nature of their antiviral activity leads to a greater chance of being effective against newly surfaced viruses, and identifying antiviral drugs from existing BSAs is an attractive strategy for responding to epidemics such as the mpox outbreaks (31). Through screening our BSA library, we identified 12 drug candidates which are comparable or more potent that the positive control cidofovir in inhibiting MPXV replication. We here prioritize safe-in-human BSAs, which have been used in the clinic or have passed phase I clinical trials, and thus these candidates can be expeditiously proceeded into clinical testing for the new indication (32).

We found clofarabine, an FDA-approved drug for treating relapsed or refractory acute lymphoblastic leukemia in children (23), as a potent inhibitor of MPXV. A study reported an EC50 value of 0.92 µM for clofarabine in inhibiting the modified vaccinia Ankara strain, but they noted the cytotoxic effects of this drug (33). In contrast, we observed that clofarabine at 1 µM exerts minimal or no toxic effects on intestinal or skin organoids. This discrepancy may be explained by the fact that their (cancer) cell line models are very sensitive to this anti-cancer drug (33). In line with our observations, previous pharmacokinetics and pharmacodynamics studies have reported the median plasma clofarabine level of 1.5 µM (range, 0.42-3.2 µM), and the median cellular concentration of 19 µM (range, 3-52 µM) (34). In this study, we determined the IC50 value of clofarabine in inhibiting MPXV replication as low as about 0.1 µM, and performed extensive validation using a concentration of 1 µM. Thus, these effective concentrations are highly clinical relevant and achievable in treated patients (34, 35), We further validated its anti-MPXV activity in human skin organoids (24). However, the antiviral activity appears more potent in intestinal compared to skin organoids. This may be attributed to relatively low bioavailability of the treated drug in the skin organoid model (24). Current clinical application of clofarabine is administered via intravenous infusion, which in general has high bioavailability, but topical formulations may be considered to further enhance skin bioavailability in case of treating mpox skin lesions (36).

As a purine nucleoside analog, clofarabine has been shown to inhibit HIV replication through both host- and virus-targeted mechanisms, inhibiting cellular nucleotide synthesis and directly inhibiting the DNA polymerase activity of HIV reverse transcriptase (37). Although the anti-MPXV mechanisms of clofarabine remain to be investigated, they are likely distinct from the mode-of-action of tecovirimat that directly targets the viral protein VP37 to prevent infectious virus production (13). Considering their complementarity, it would be interesting to assess the combination of clofarabine with tecovirimat to potentially achieve synergistic antiviral activity and to prevent drug resistance development (38).

The epidemiological and clinical features of mpox substantially depend on the viral (sub-)clades and the exact context. The 2022-2023 global outbreak of clade IIb primarily affected men who have sex with men through sexual network (9). Historically, the infection of clade I MPXV was thought to be more pathogenic (39), but its transmission was unrelated to sexual activity. However, the ongoing outbreak of clade I in DRC has documented the presence of sexual transmission (40). Any population can get infected with MPXV, but vulnerable populations such as children, pregnant women and immunosuppressed individuals (e.g. those infected with HIV) are at higher risk of developing severe complications (41). Currently, the clade Ia and Ib strains are co-circulating in Africa (3), whereas the new variant clade Ib harbors novel mutations (4).

Of note, there are some limitations in this study. First, the distinct susceptibilities of differentiated cell types from intestinal organoids to MPXV infection require further mechanistic understanding. Second, our models support robust infections of the different subclades of MPXV, but still not yet fully recapitulate their (differential) disease manifestations. Third, the discovered anti-MPXV drug clofarabine requires further evaluation before proceeding into clinical testing for treating mpox, and the anti-MPXV mechanisms should be investigated in future research. Fourth, our organoid models mainly recapitulate MPXV acute infection and short-term antiviral treatment. However, prolonged mpox may occur in peculiar cases (e.g. with advanced HIV) (42), and longer-term treatment is likely required to benefit these patients. Therefore, future research should also dedicate to the development of experimental models recapitulating MPXV persistent infection and testing long-term treatment, as well as monitoring potential drug resistance emergence. Finally, we did not test clade IIa, but this subclade is mainly restricted to a few countries in West Africa, has low rates of human-to-human transmission, and has not recently been a significant cause of mpox epidemics (43). However, in 2024, clade IIa cases were reported in Côte d’Ivoire, Guinea, and Liberia, marking the first evidence of sustained community transmission (44), which should attract more attention for future research.

In summary, this study successfully modeled the infections of clade Ia, Ib and IIb isolates in human intestinal organoids, and demonstrated the potent antiviral activity of clofarabine across these three subclades. Thus, our innovative experimental model and the antiviral drug discovery pipeline bear major implications in responding to the current mpox global health emergency, and sustaining epidemic poxvirus preparedness.

## Materials and methods

### Intestinal organoids culture

Human primary intestinal organoids were isolated and cultured as we previously described (21, 45). The use of human intestinal tissue for research purpose including culturing into organoids was approved by the Medical Ethical Council of the Erasmus MC, and informed consent was given (MEC-2021-0432; MEC-2023-0629). These organoids were cultured in organoid expansion medium (OEM), based on advanced DMEM/F12 (Invitrogen), supplemented with 1% penicillin/streptomycin (Life Technologies), 10 mM HEPES, 1xGlutamax, 1 mM N2, 1 mM B27 (all from Invitrogen), 1 μM N-acetylcysteine (Sigma) and the following growth factors: 50 ng/L mouse epidermal growth factor (mEGF), 50% Wnt3a-conditioned medium (WCM) and 10% noggin-conditioned medium (NCM), 20% Rspo1-conditioned medium, 10 μM nicotinamide (Sigma), 10 nM gastrin (Sigma), 500 nM A83–01 (Tocris) and 10 μM SB202190 (Sigma). The medium was refreshed every 2-3 days, and organoids were passaged 1:3 every 5–7 days.

### Differentiation of organoids

Organoids were further differentiated towards different cell types by using the respective differentiation medium. Briefly, organoids were cultured in OEM (without WCM) supplemented with 2 µM IWP-2 (Sigma) for enterocytes differentiation; OEM (without WCM) supplemented with 2 µM IWP-2 and 10 µM DAPT (MedChemExpress) for goblet cells differentiation; OEM (without WCM) supplemented with 2 µM IWP-2, 10 µM DAPT and 5 µM Gefitinib (Sigma) for enteroendocrine cell differentiation. The differentiation medium was refreshed every 2-3 days and expected cell types were achieved after 5 days differentiation culture.

### Virus inoculation

Intestinal organoids were mechanically fragmented and then exposed to MPXV viral particles for 1 hour at 37°C. Every 1000 organoids were exposed to approximately 10^4 PFU viruses. To increase the infection efficacy, organoids and virus mixture were re-suspended every 20 minutes throughout the inoculation period. Subsequently, fragmented organoids underwent centrifugation at 300 g for 5 minutes at 4°C, and the supernatant was discarded. Then organoids were thoroughly washed three times with advanced DMEM/F12 to remove residual viruses. After infection, the organoids were embedded in Matrigel and cultured in OEM.

### Organoid cells cultured in trans-well system

Intestinal organoids were digested into single cells by TrypLE Express, and approximately 2.4x10^4 cells were then seeded in each trans-well insert (pre-coated with 10-fold diluted Matrigel), with 200 uL and 450 uL OEM supplemented in the apical and basolateral compartment respectively. After cells growing into full confluence, MPXV viral particles were inoculated from apical side of the insert.

### AlamarBlue assay

Intestinal organoids were cultured in organoid expansion medium. Mature skin organoids were cultured in organoid maturation medium as one organoid per well. After removing culture supernatant, intestinal organoids or skin organoids were incubated with a 1:20 dilution of AlamarBlue regent (Invitrogen, DAL1100) in culture medium for 2 hours at 37°C. Subsequently, 100 μL medium was collected to assess cell metabolic activity, with each sample being measured in duplicate. Absorbance measurements were obtained using a fluorescence plate reader (CytoFluor Series 4000, PerSeptive Borganoidsystems) at an excitation wavelength of 530/25 nm and an emission wavelength of 590/35 nm.

### DNA extraction and qPCR detection

Total DNA was purified from infected organoids or culture medium using Macherey-Nagel NucleoSpin DNA Kit (Bioke, Netherlands) and quantified by Nanodrop ND-1000 (Wilmington, USA). Viral DNA levels were quantified by SYBR Green-based qRT-PCR (Applied Biosystems SYBR Green PCR Master Mix; Thermo Fisher Scientific Life Sciences) with the StepOnePlus System (Thermo Fisher Scientific Life Sciences). MPXV viral DNA was calculated by previously generated formula “y=-0.3095x+15.387”. The primers used in this study were provided in Supplementary Table 1.

### Plaque assay

Organoids were collected and stored in 1 mL serum-free advanced DMEM/F12 medium (with 1xGlutaMAX, 1 M Hepes and 1% penicillin/streptomycin), and centrifuged after 3 cycles of freezing and thawing to collect clear cell lysates as intracellular viruses. Cleared supernatants from culture medium were used as extracellular viruses. Confluent Vero cells in 12-well plates were washed once with PBS before virus inoculation. Next, Vero cells were overlaid with 1.2% avicel in serum-free advanced DMEM/F12 medium (with 1xGlutaMAX, 1 M Hepes and 1% penicillin/streptomycin) containing ten-fold serial dilutions of samples. Plates were incubated 4 days at 37°C and then fixed by 4% paraformaldehyde (PFA) and stained with 0.1% crystal violet. Plaques were quantified as PFU/mL. Data were presented as the common logarithm, and the samples with undetectable plaque were arbitrarily denoted with a value “1”.

### Immunofluorescence staining and confocal imaging

Organoids were fixed with 4% paraformaldehyde (PFA) for 15 min. Then the samples were gently rinsed 3 times with PBS, followed by permeabilizing with PBS containing 0.2% (vol/vol) Triton X-100 for 10 min. Next, samples were twice rinsed with PBS for 5 min, followed by incubation with blocking solution (5% donkey serum, 1% bovine serum albumin, 0.2% Triton X-100 in PBS) at room temperature for 1 hour. Primary antibodies diluted in blocking solution were subsequently incubated with samples at 4°C overnight. Samples were then washed 3 times for 5 min each in PBS prior to 1 hour incubation with 1:1000 dilutions of the secondary antibodies Nuclei were stained with DAPI (4, 6-diamidino-2-phenylindole; Invitrogen). At last, stained samples were visualized using a Leica SP5 confocal microscope to analyze the stained cellular structures. Antibodies used in this study were listed in the Supplementary Table 2.

### Genome-wide RNA sequencing and data analysis

Intestinal organoids cultured from human ileum tissue were infected with MPXV clade IIb (NL001 strain) viral particles as above description. Infected organoids were harvested at 1, 24, 48 and 96 hours post-infection. Infected organoids with 1 μM clofarabine treatment starting from 1 hour till 96 hours was set up as a treatment group. In parallel, un-infected organoids were cultured under same conditions for 96 hours as negative controls. Total RNA was isolated using the MachereyNagel NucleoSpin RNA II Kit (Bioke, Netherlands) and quantified with the Bioanalyzer RNA 6000 Picochip. Afterwards, RNA sequencing was conducted by Novogene using a paired-end 150 bp (PE 150) sequencing strategy. RNA sequencing data are publicly available at Data Archiving and Networked Services (DANS) (https://doi.org/10.17026/LS/MDIBMO).

### Statistics

Statistical analysis was performed using GraphPad Prism8 statistics software (GraphPad, San Diego, USA). All data are presented as mean ± standard error of the mean (s.e.m.). Comparison between two groups was analysed by Mann–Whitney U test. Asterisks indicated the degree of significant differences compared with the controls (*, p < 0.05; **, p < 0.01).

## Data Availability

All data produced in the present work are contained in the manuscript and supplementary file.

https://doi.org/10.17026/LS/MDIBMO

## Acknowledgement

We thank Dr. Daniel Bourquain from Robert Koch Institute, Germany, for providing the clade Ia MPXV isolate, Dr. Sara Byfors from Public Health Agency of Sweden for providing the clade Ib MPXV isolate. This work was supported by a VIDI personal grant (No. 91719300) from the Netherlands Organisation for Health Research and Development (ZonMw) to Q.P; a Young Investigator Grant from Erasmus MC-University Medical Center and a pandemic preparedness grant (No. 10710032310013) from ZonMw to P.L. A.N.D.N and A.M.G. were supported by the National Council for Scientific and Technological Development, Brazil (CNPQ #445484/2023-3). K.R. was supported by the Novo Nordisk Foundation, Denmark (grant number NNF21CC0073729), and K.R. is Chargé de Recherche at the Institut National de la Santé et de la Recherche Médicale (INSERM).

