## Supplementary information for "Primary human intestinal organoids recapitulate enteric infection of monkeypox virus and enable scalable drug discovery"

*Li et al.*

Figure S1

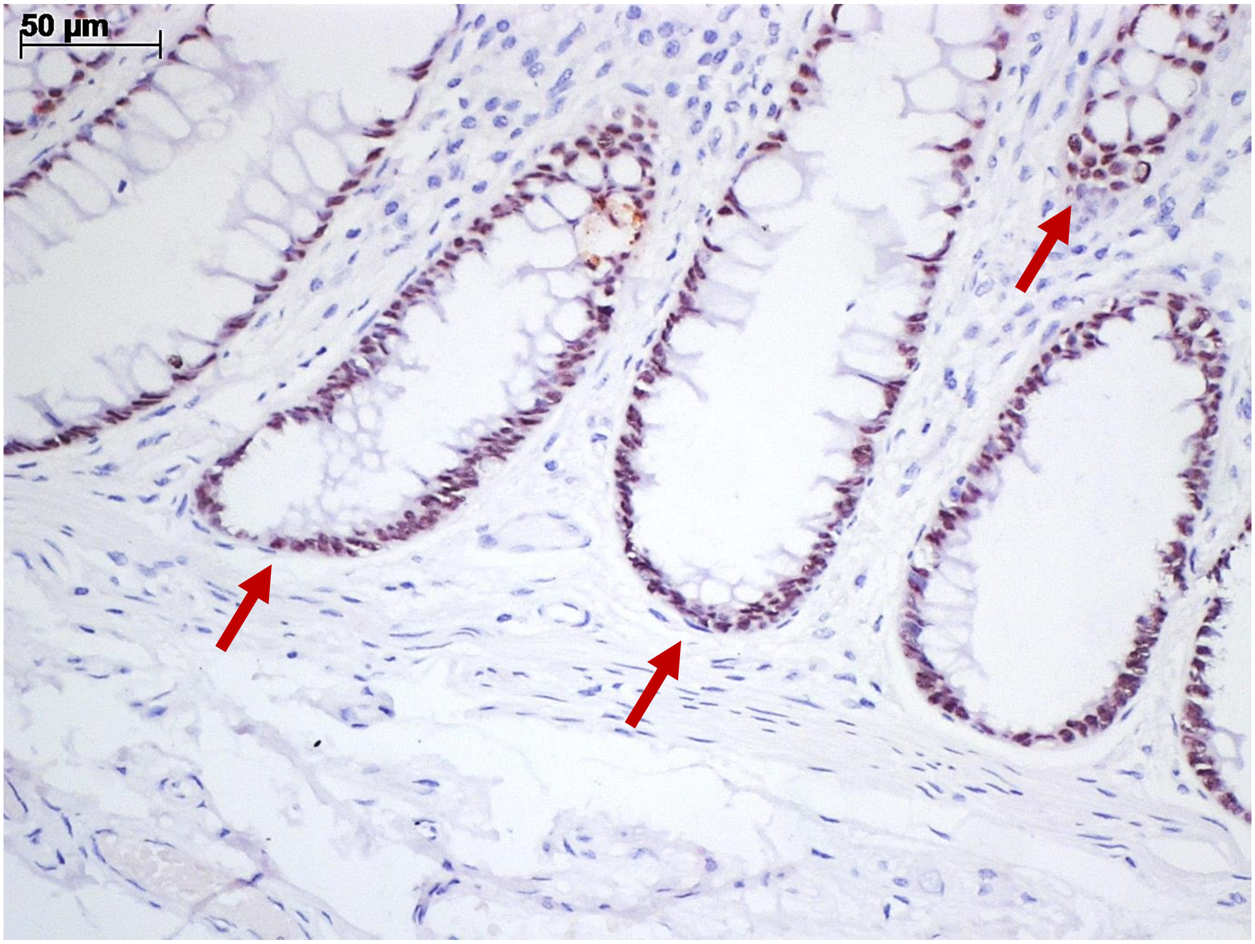


Figure S1. IHC staining of anti-CDX-2 and anti-Vaccinia in normal intestinal tissue (uninfected human case). CDX-2 positive staining displayed as nuclei of intestinal epithelium lining colonic villi and crypts (red arrow). Anti-Vaccinia was negative.

Figure S2

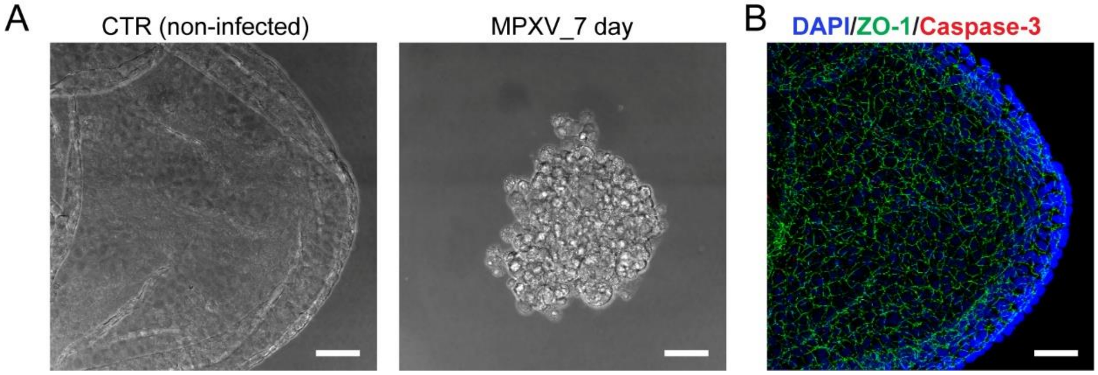


Figure S2. Morphology of MPXV infection in intestinal organoids. (A) Bright field images of non-infected organoids and MPXV-infected organoids after 7 days of infection. (B) Immunostaining of non-infected organoids (DAPI, nucleus; ZO-1, tight junction; Caspase-3, cell death).

Figure S3

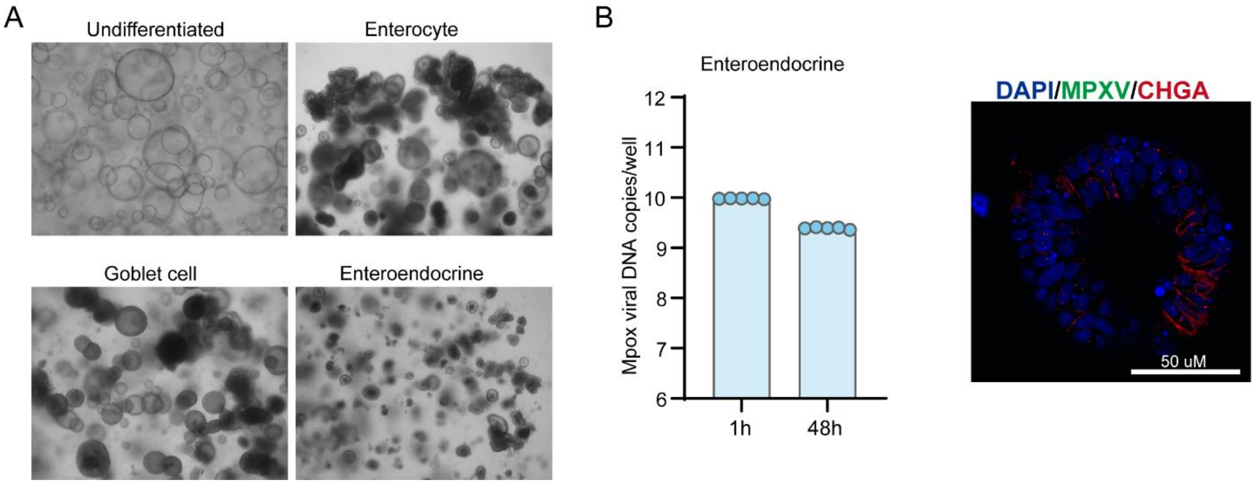


Figure S3. Modelling MPXV infection in differentiated intestinal organoids. (A) Morphology of organoids differentiating towards enterocytes-, goblet- and enteroendocrine (EEC)-phenotypes after 5-days differentiation culture. (B) QRT-PCR quantification of viral DNA level in enteroendocrine-differentiated organoids and immunostaining MPXV virions at 48 hours post-infection. CHGA (red) is the representative marker of enteroendocrine cells. MPXV (green) florescence signal was not detected in CHGA-positive organoid cells.

Figure S4

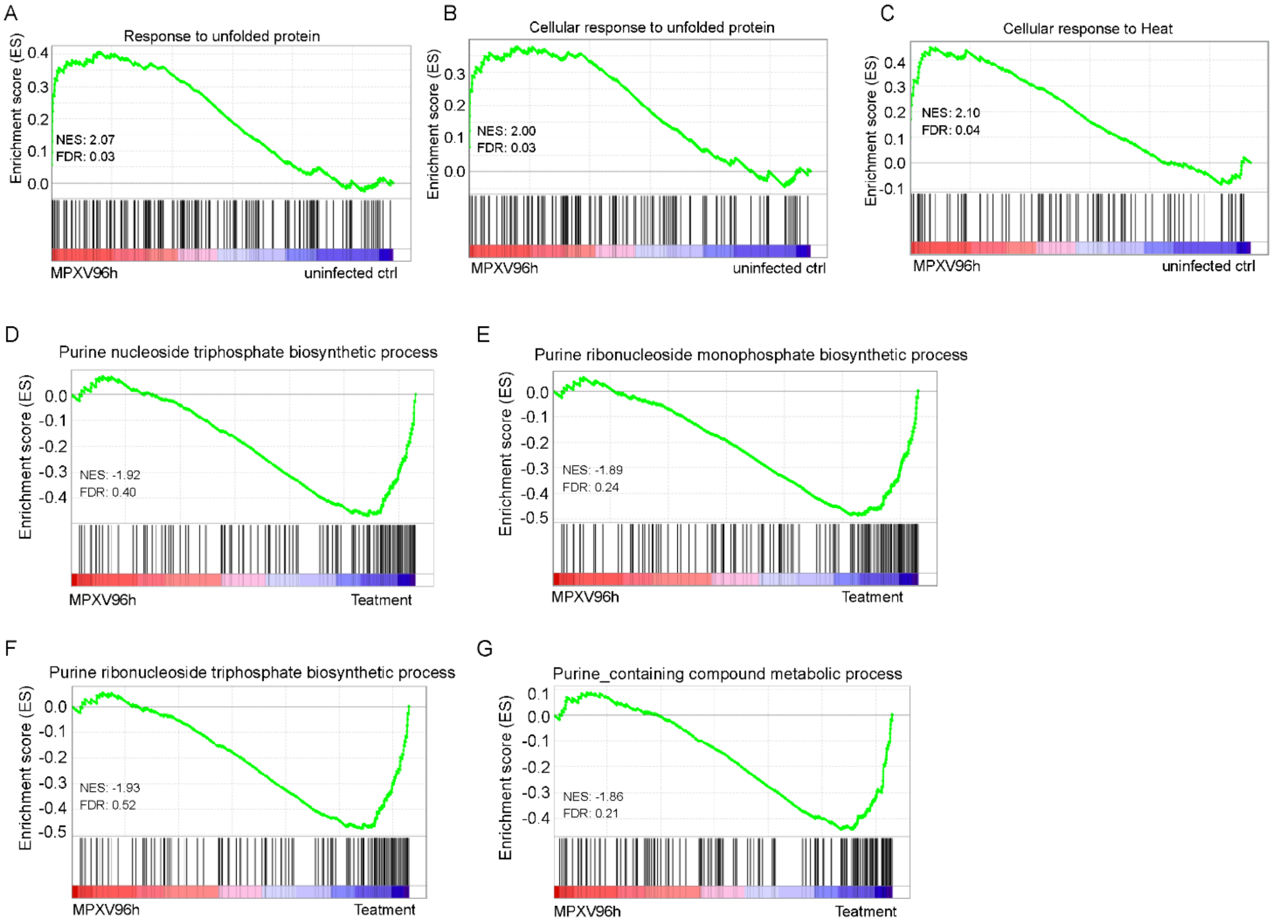


Figure S4. Gene set enrichment analysis (GSEA) of GO pathways in intestinal organoids infected with MPXV. (A-C) Enriched transcriptional signatures in organoids of 96 h.p.i comparison with uninfected organoids. (D-G) Comparison of MPXV infected organoids for 96 hours with and without clofarabine treatment showed that the transcriptional signatures of purine nucleoside/ribonucleoside associated process were inhibited.

Figure S5

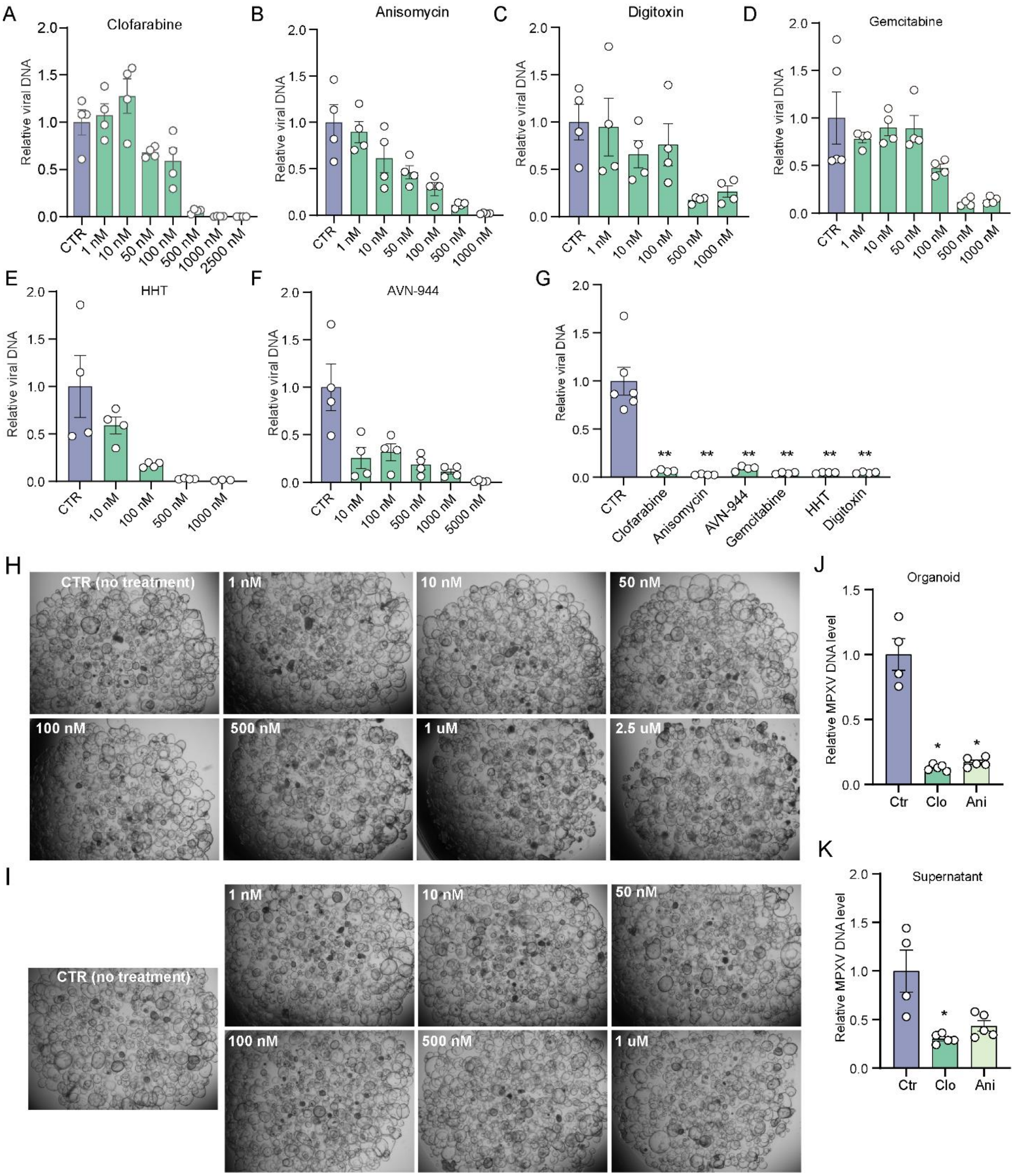


Figure S5. The antiviral effects of identified leading compounds including clofarabine (A), anisomycin (B), digitoxin (C), gemcitabine (D), homoharringtonine (HHT) (E), and AVN-944 (F) in MPXV-infected intestinal (ileum) organoids after 48 hours treatment. (G) The antiviral effects of tested compounds in human rectum organoids infected with MPXV and treated for 48 hours.

Bright field images of intestinal (ileum) organoids treated with different concentrations of clofarabine (H) and anisomycin (I) for 48 hours. Quantification of MPXV DNA level in organoids (J) and culture medium (K) after 7 days treatment with clofarabine (Clo) and anisomycin (Ani).

Supplementary Table 1. Primers used in this study.

| Gene | Sequence (5'-3') |
|---|---|
| GAPDH-F | GTCTCCTCTGACTTCAACAGCG |
| GAPDH-R | ACCACCCTGTTGCTGTAGCCAA |
| Villin-F | GCTGCTCTACACCTACCTCATC |
| Villin-R | TTCTGGTCCAGGATGACGGCTT |
| Muc-2 F | ACTCTCCACACCCAGCATCATC |
| Muc-2 R | GTGTCTCCGTATGTGCCGTTGT |
| hCHGA-F | TGACCTCAACGATGCATTTC |
| hCHGA-R | CTGTCCTGGCTCTTCTGCTC |
| MPXV-F | GGCTCTTCTATCAACCACA |
| MPXV-R | AGTCATTATCTCCTCCTCCA |

Supplementary Table 2. Antibodies used in this study.

| Antibody | Company & Catalogue number |
|---|---|
| Rabbit polyclonal anti-Vaccinia virus Lister Strain (FITC) | Abbexa, abx023199 |
| Rabbit polyclonal anti-Vaccinia virus Lister Strain | Abbexa, abx023200 |
| Anti-CDX2 antibody, rabbit | Abcam, AB76541 |
| Anti-SOX9 antibody, rabbit | Abcam, AB185966 |
| Anti-Ki67 antibody, rabbit | Abcam, AB15580 |
| Anti-ZO-1 antibody, | Life technologies, MA339100A488 |
| Anti-EpCAM, rabbit | Abcam, ab71916 |
| Anti-Villin, mouse | Santa Cruz Biotechnology, sc-66022 |
| Anti-Muc2, mouse | Santa Cruz Biotechnology, sc-59859 |
| Anti-CHGA, mouse | Santa Cruz Biotechnology, sc-393941 |
| Alexa Fluor 488 Goat anti-Rabbit antibody | ThermoFisher Scientific, A32731 |
| Alexa Fluor 594 Goat anti-Mouse antibody | ThermoFisher Scientific, A32742 |
